# Global left ventricular wall thickness by coronary computed tomography angiography – derivation, validation, and normal reference values

**DOI:** 10.1101/2025.04.27.25326520

**Authors:** Raquel Themudo, Andreas Fridén, Magnus Lundin, Einar Heiberg, Tomas Jernberg, Martin Ugander

## Abstract

**Aim:** Left ventricular hypertrophy (LVH) is a prognostic marker in the assessment of cardiovascular disease. Left ventricular mean global wall thickness (GT) can be measured using cardiovascular magnetic resonance, and has been shown to improve diagnostic and prognostic performance beyond left ventricular mass (LVM). The aim of this study was to use coronary computed tomography angiography (CTA) to derive and validate a formula to estimate GT from coronary CTA, and to determine normal reference values.

**Methods and results:** This was a retrospective cohort study among subjects aged 50-64 years randomly selected from the general population as part of the Swedish CArdioPulmonary bioImage Study (SCAPIS). GT, LVM, and left ventricular mid-diastolic volume (LVMDV) were measured by coronary CTA. A formula for estimating GT using only LVM and LVMDV was derived and validated. The study included 414 subjects (age 59±4 years, 67% male). Calculated GT was derived in a derivation subset (n=207) as GT[mm]=1.55xLVM[g]^0.77xLVMDV[ml]^-0.41, and agreed with measured GT in a separate validation subset (n=207, R^2^=0.97, p<0.001, bias 0.01±0.23 mm). Among a subset of healthy subjects, calculated GT for females (n=27) was 7.0±0.9 mm (normal range 5.2-8.8 mm), and for males (n=20) was 8.9±1.3 mm (normal range 6.4-11.4 mm). Systolic blood pressure, age and coronary artery calcium score were all weakly correlated with GT (R^2^=0.03–0.11, p<0.05 for all).

**Conclusion:** GT from coronary CTA can be calculated using only LVM and LVMDV with excellent accuracy and precision, and sex-specific normal reference values in healthy subjects are presented for use in the clinical evaluation of LVH.

## Introduction

Left ventricular hypertrophy (LVH) is defined as an increase in left ventricular mass, either due to an increase in ventricular wall thickness or due to an enlargement of the left ventricular cavity, or both.^1^ The pathophysiologic process underlying myocardial hypertrophy is not completely understood, but it is thought to be influenced by mechanical factors such as the hemodynamic overload activating myocardial growth, as well as non-mechanical factors, such as genetic, ethnic, environmental and hormonal factors, contributing to the development of left ventricle hypertrophy.^2–4^ LVH is proven to be a cardiovascular risk factor beyond other conventional risk factors in population based-studies. ^5–7^ Importantly, there is evidence that regression of LVH in patients with hypertension is associated with a reduction in cardiovascular events.^8^

Coronary computed tomography angiography (CTA) is an imaging technique commonly used to diagnose or exclude coronary artery disease in patients with low to intermediate risk for coronary artery disease. Coronary CTA is a first-line investigation for the evaluation of stable coronary artery disease and is frequently used to investigate patients who have equivocal findings in other imaging modalities.^9–11^ In addition to assessment of coronary arteries, coronary CTA can provide simultaneous information about the morphology and function of the myocardium, including left ventricular mass and left ventricular volume. Coronary CTA can thus be used as an alternative to cardiovascular magnetic resonance (CMR) in the evaluation of patients with LVH who have contraindications to undergo CMR.^12^ Also, patients undergoing assessment for coronary stenosis by coronary CTA may also benefit from risk assessment regarding presence or absence of LVH. Standard coronary CTA acquisitions include, most commonly, only images acquired in mid-diastole, which will necessarily yield a greater wall thickness size compared to images acquired in end-diastole, which is the standard for echocardiography and CMR.^12^ Therefore, normal reference values for left ventricular mass and wall thickness on standard coronary CTA at mid-diastole in healthy individuals need to be determined.

A new method to accurately and precisely estimate the so called left ventricular global wall thickness (GT) based on geometrical assumptions of the left ventricle shape using measurements of only left ventricular end-diastolic volume (LVEDV) and left ventricular mass (LVM) using CMR has recently been developed.^13^ GT provides an intuitive measure of the balance between left ventricular volume and mass, where the calculated GT has been proven to have excellent agreement with measured GT of the left ventricle by CMR.^13^ Moreover, GT has been proven to be a strong independent predictor of death and hospitalization for heart failure in patients with otherwise normal findings on CMR. GT of the left ventricle indexed to body surface area (GTi) has also been demonstrated to have prognostic utility primarily among patients with normal LVEDV. However, GT has not been evaluated by coronary CTA. Therefore, the aim of this study was to derive and validate a GT formula for coronary CTA, and to determine normal reference values, in order to more accurately assess LVH using coronary CTA.

## Methods

### Patient data

The Swedish CArdioPulmonary bioImage Study (SCAPIS) recruited subjects at six University hospitals in Sweden. The study included 30,154 participants in the age group of 50 to 64 years, randomly selected from the general population in Sweden with a participation rate of 50.3%.^14^ Participants were included between the years 2013 and 2018 and were subject to comprehensive examination and testing, including coronary CTA imaging. For the purposes of this study, the SCAPIS registry provided access to data from 500 subjects from the Stockholm recruitment site who had undergone coronary CTA. The study was approved by the Regional Ethics Board in Stockholm, Sweden, with the registration number 2018/2605-31/1 2019-00274, and all participants provided written informed consent.

Exclusion criteria for all patients included cardiovascular disease, defined as prior myocardial infarction or revascularization, valvular heart disease, heart failure, congenital heart disease, heart transplant, or prior cardiac surgery. Other exclusion criteria included systemic disease such as diabetes mellitus, chronic kidney disease (glomerular filtration rate < 30 ml/min/1.73 m^2^), connective tissue disease, cardiomyopathy, or pathological findings on ECG by automatic interpretation. No exclusion was performed based on pre-existing medication. Heart rate greater than 100 beats/minute during image acquisition or acquired images outside of the 66-76% cardiac cycle window also lead do exclusion. A flowchart describing the subject selection process is presented in Fig. 1.

**Figure 1.**
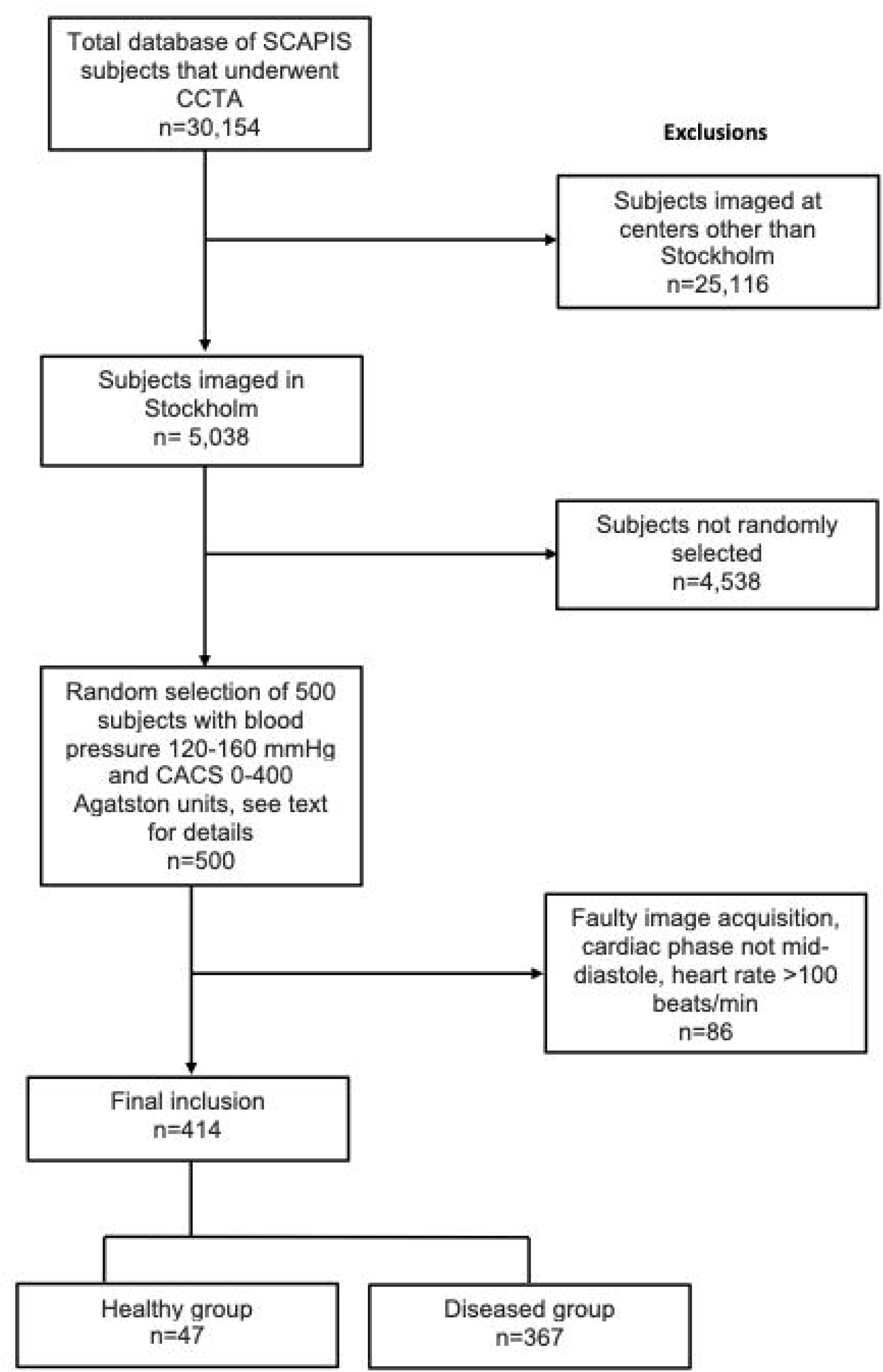
Flowchart of the study population selection. From the original 30,154 volunteers in the SCAPIS study nationally, 5,038 were recruited in Stockholm. From these subjects, 500 were randomly selected from a subgroup with systolic blood pressure between 120-160 mmHg and Agatston score 0-400 Agatston units (AU). In the chosen group of 500, exclusion criteria were image acquisition window outside of the 66-76% RR interval, heart rate >100 beats/minute or suboptimal image quality (n=86; 82 subjects due to suboptimal image quality, 2 due to heart rate >100 beats/minute, 1 due to image acquisition outside of the 66-76% RR interval and 1 subject due to dwarfism). The study population was then divided into a healthy group if systolic blood pressure (SBP) <140 mmHg and Agatston score 0 (n=47). The remaining subjects (SBP >=140 mmHg and Agatston score ≠ 0) were categorized as the diseased group (n=367).

In order to obtain a variation in left ventricle wall thickness, the five hundred subjects were primarily randomly selected from the SCAPIS Stockholm region cohort in order to obtain five groups with different systolic blood pressure, divided by 10 mmHg increments starting at 120 mmHg, and then further divided according to the following Agatston score: 0, 1-99, and 100-400 AU. The final study population (n=414, 33% female) was divided into the Healthy Group (n=47, 60% female, systolic blood pressure (SBP) <140 mmHg, Agatston score 0 AU) and Diseased Group (n=367, 29% female), see Table 1. The derivation and validation of the formula for GT was based on the final study population, divided into equal parts of females and males in the derivation subset (n = 207), and a separate validation subset (n=207).

**Table 1.**
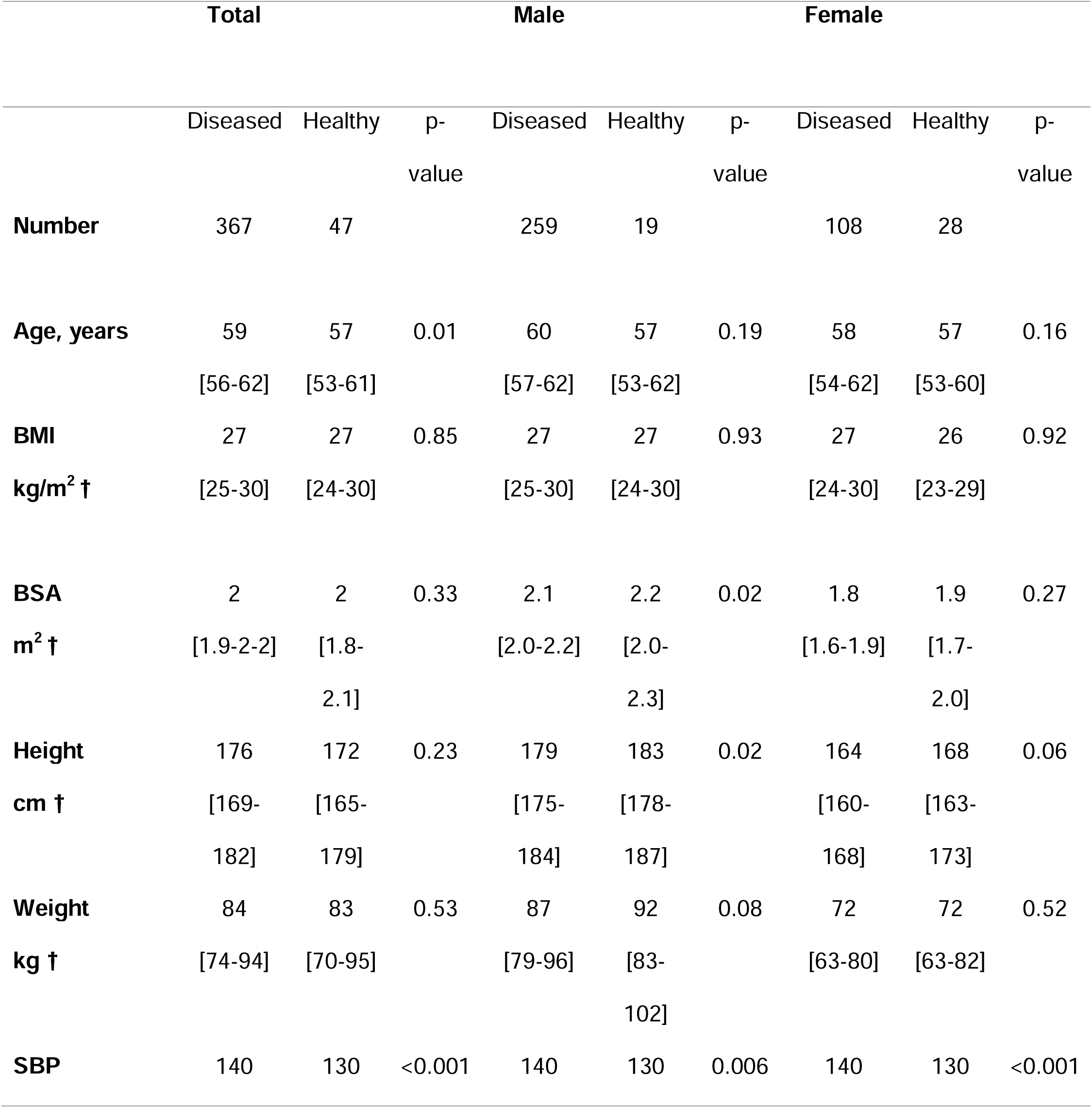
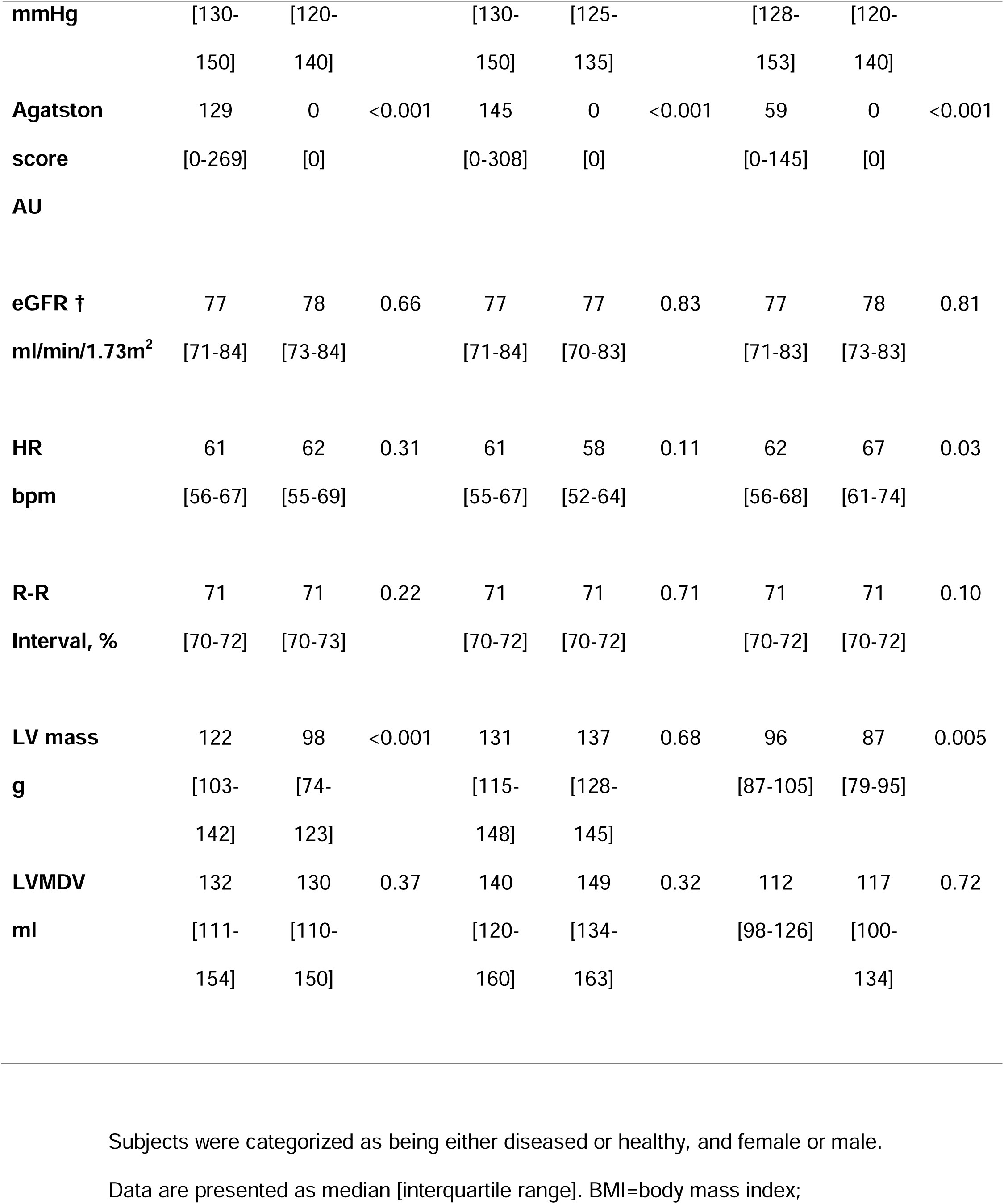
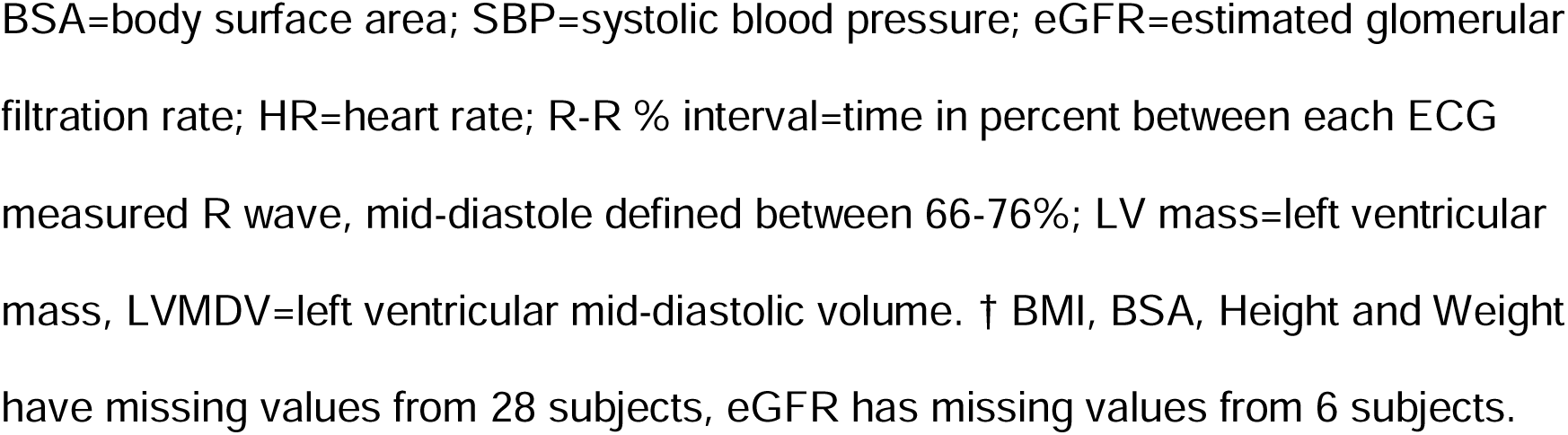
Baseline characteristics of the diseased and healthy study population according to gender (n=414).

### CT Image Acquisition

CT imaging was performed with a second generation dual-source Stellar detector (Somatom Definition Flash; Siemens Medical Solution, Forchheim, Germany). A non-contrast scan over the heart was first acquired with prospective ECG triggering and 120 kVp and reconstructed to 3-mm thick slices in order to measure coronary calcium score. Coronary CTA images were then acquired with an automated tube current at 100-120kVp and the use of intravenous contrast medium (iohexol, 350 mg I/mL, Omnipaque, GE Healthcare, Stockholm, Sweden). An intravenous or oral beta-blocker was used to reduce heart rate with a target heart rate of less than 60 beats/minute. Additionally, sublingual nitroglycerin (4 mg/dose; G. Pohl-Boskamp GmbH & Co. KG, Hohenlockstedt, Germany) was also used to induce vasodilatation.

### Image analysis

- Measurements of LV-mid diastolic volume, mass and wall thickness All image analysis was performed using the software Segment CT (Medviso AB, version 3.0 R7990, Lund, Sweden). Thin axial slices from the coronary angiography series were reformatted into 10 mm thick short-axis slices of the left ventricle, see Fig. 2. The software performed semi-automatic tracing of endo- and epicardial borders, and then manual adjustments were performed. GT was measured automatically at 24 measurement points at equal distance around the circumference of the left ventricle for every 10 mm thick slice (Fig. 3). Regions with wall thickness less than 2 mm were excluded. The mean wall thickness per slice was weighted according to the slice circumference and averaged across the whole left ventricle. Measurements of left ventricular mass and volume were also noted.

**Figure 2.**
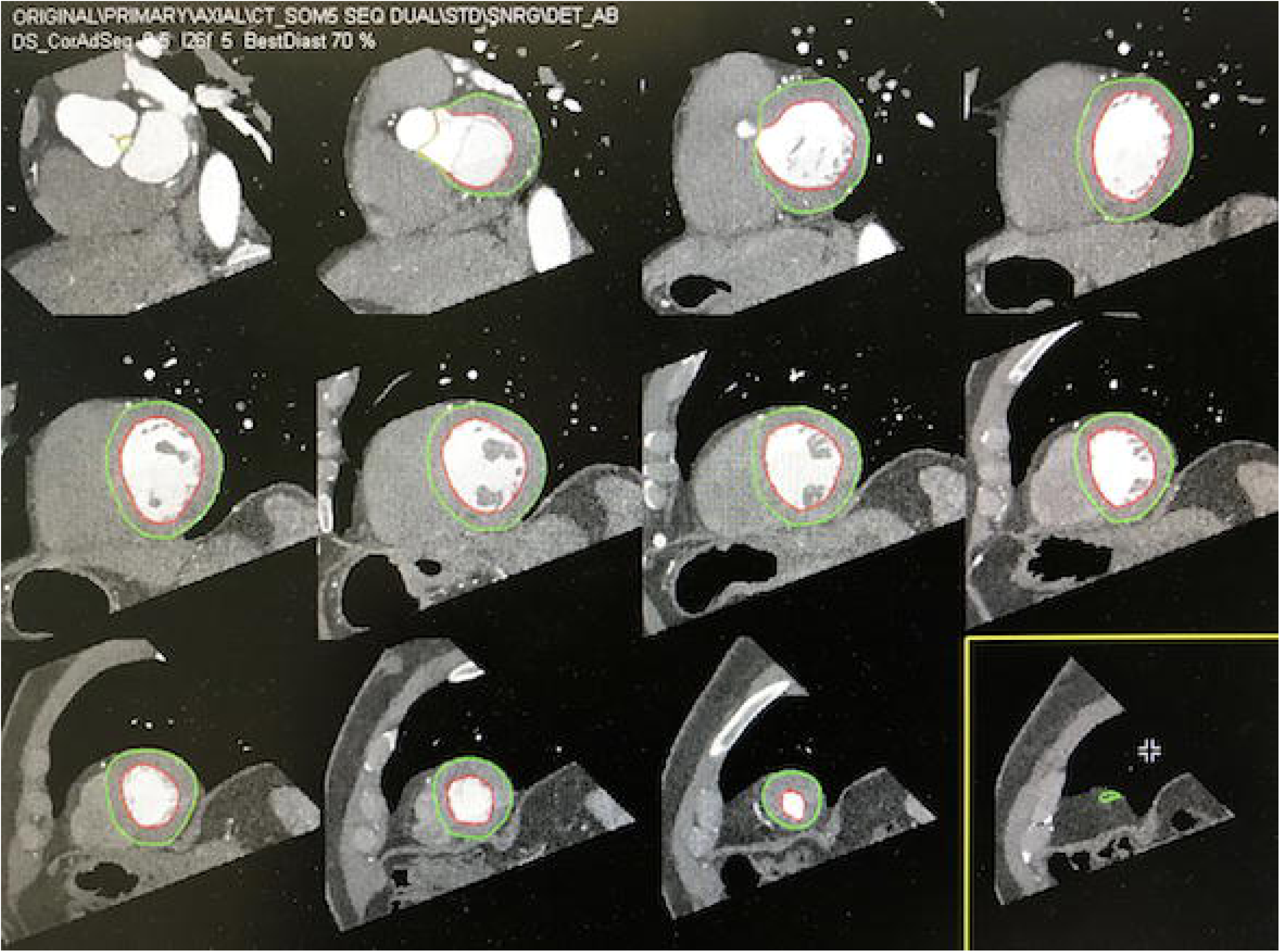
Segmentation of left ventricular short-axis slices from coronary CTA. Short-axis view covering the left ventricle from base to apex with a slice thickness of 10 mm. The LV endocardial (red line) and epicardial (green line) borders have been automatically traced and manually adjusted if needed. LV mid-diastolic volume and mass were calculated from this segmentation. Papillary muscles and myocardial trabeculation were included in the blood pool volume.

**Figure 3.**
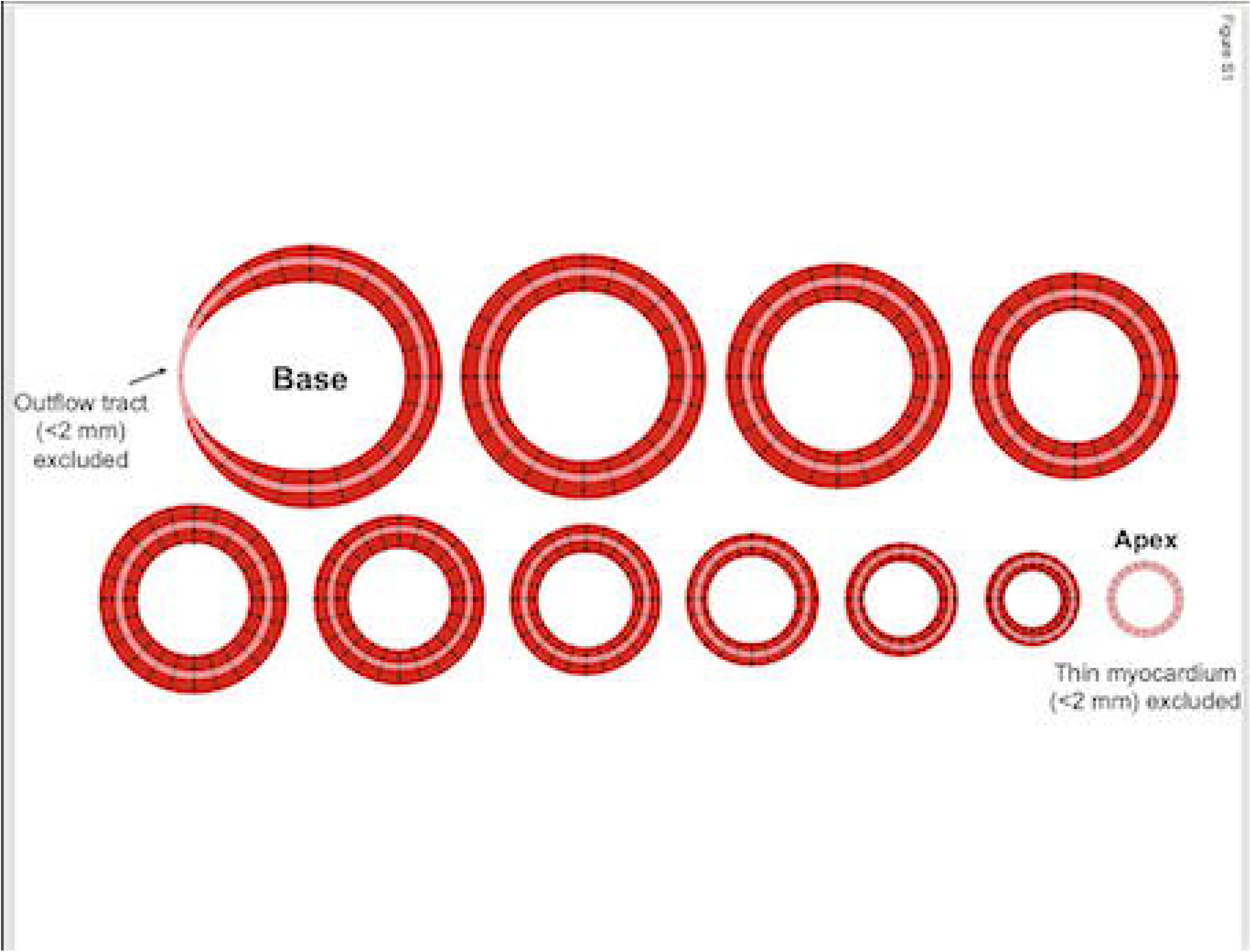
Method of GT measurement. After endo- and epicardial borders tracing, a plugin from the Segment CT software was used in order to measure global wall thickness in 24 positions for each of the short-axis slices. The distance between the endocardial and epicardial borders at end-diastole was measured at 24 evenly distributed positions (shown as dashed black lines) around the circumference of all short-axis slices of a full LV short-axis stack from base to apex. Basal sections with a wall thickness of less than 2 mm in the LV outflow tract and the apex were excluded. The mean thickness for each individual short-axis slice was multiplied by the midmural circumference of that slice (pink circle), these were summed and then divided by the sum of the midmural circumference for all slices to yield the GT. Reproduced from Lundin M, et al^13^, with permission. https://www.nature.com/articles/s41598-023-48173-7#rightslink

### Estimating GT from LV volume and mass

A mathematical equation developed for use with CMR was used to estimate GT from mass and volume by coronary CTA as:

GT = A + B • LVM^x^ • LVMDV^y^

LVMDV: left ventricular mid-diastolic volume.

The constant A, coefficient B, and exponential coefficients x and y, that gave the best fit to measured GT, were calculated using in-house developed software implemented in MATLAB (R2016, Math, Natick, Massachusetts, USA). This method has been described in greater detail previously.^13^ The derivation of the equation for estimating GT was performed using the derivation subset (n=207). Calculated GT was then validated in the validation subset. The validated formula was then used to calculate GT for the healthy group of females (n=27) and males (n=20).

### Statistical Analysis

Statistical analysis was performed in the open-source software R (R Core Team, Vienna, Austria). Normality of distribution was tested using the Shapiro-Wilks and Kolmogorov-Smirnov tests. Non-normally distributed data was presented as median [interquartile range], and differences between groups were tested using the Wilcoxon test. Sex-specific normal values of GT were presented as the range between mean-1.96SD to mean+1.96SD. Linear regression analysis was performed and Pearson’s or Spearman’s correlation coefficients were calculated as appropriate. Bland-Altman analysis was also performed. A p-value less than 0.05 was considered statistically significant.

## Results

### Derivation, validation and normal ranges for GT

The patient characteristics for the derivation and validation groups are presented in Table 1.

Using the derivation subset (n=207), the optimized equation for the calculated global wall thickness (GT) was found to be:

GT [mm] = -0.0002 + 1.5474 · LVM[g]^0.7694^ · LVMDVI[ml]^-0.4107^

where GT is the global wall thickness in mm, LVM is left ventricular mass in grams, and LVMDV is left ventricular mid-diastolic volume in milliliters. For the derivation subset, the model had an expectedly high agreement between calculated and measured GT (R^2^=0.97, p<0.001, bias 0.00±0.23 mm, top row figure 4). When applied to the separate validation subset of the cohort (n=207), the model performance was preserved (R^2^=0.97, p<0.001, bias -0.01±0.25 mm, bottom row figure 4).

**Figure 4.**
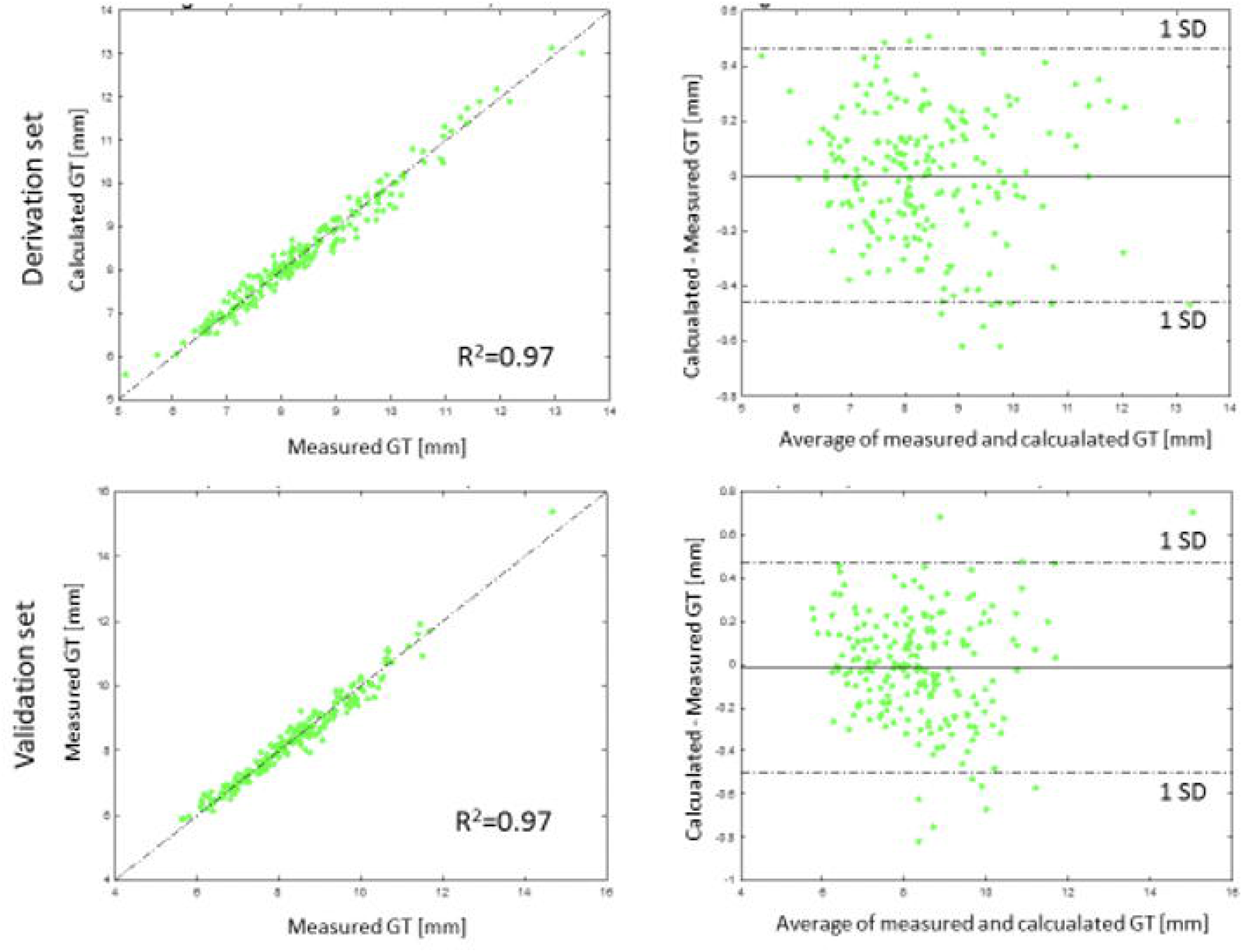
Linear regression and Bland-Altman plots comparing calculated GT and measured GT in the derivation and in the validation subgroups. Top left: Correlation plot for the derivation subset (n=207), R^2^=0.97, (p<0.001, bias 0.00±0.23 mm), identity line shown dashed. Top right: Bland–Altman plot for the derivation subset of the cohort. Solid line shows a mean difference and dashed lines show ±1.96 standard deviations. Bottom left: Correlation plot for the separate validation subset (n=207), R^2^=0.97, (p<0.001, bias -0.01±0.25 mm), identity line shown dashed. Bottom right: Bland–Altman plot for the validation subset of the cohort. The solid line shows mean difference and dashed lines show ±1.96 standard deviations. GT=global wall thickness.

The calculated GT in healthy volunteers (n=47, 60% female) was 7.0±0.9 mm for females and 8.9±1.3 mm for males. This corresponds to a GT with a normal reference range of 5.2–8.8 mm for females and 6.4–11.4 mm for males.

When GT was indexed for body surface area (GTi), the values were 3.9±0.6 mm/m^2^ for females and 4.1±0.6 mm/m^2^ for males, corresponding to normal ranges of 2.7-5.1 mm/m^2^ for females and 2.9-5.3 mm/m^2^ for males. LVMi for the healthy volunteers was 48±6 g/m^2^ for females and 63±11 g/m^2^ for males and LVMDVi was 62±6 ml/m^2^ for females and 69±10 ml/m^2^ for males.

Calculated GT with the coronary CTA formula presented herein in the whole cohort (n=414) (8.2±1.3 mm) agreed with calculated GT with the CMR formula, previously described as

GT [mm] = 0.05 + 1.6 · LVM[g]^0.84^ · LVMDV[ml]^-0.49^ ^13^, (8.1±1.4 mm, R^2^=0.9997, p<0.001, bias 0.09±0.15mm).

### Associations of GT with age, blood pressure, and coronary artery calcium score

The association between GT and blood pressure, age, and Agatston score was evaluated in the whole study population (n=414) divided by sex, table 2.

**Table 2.**
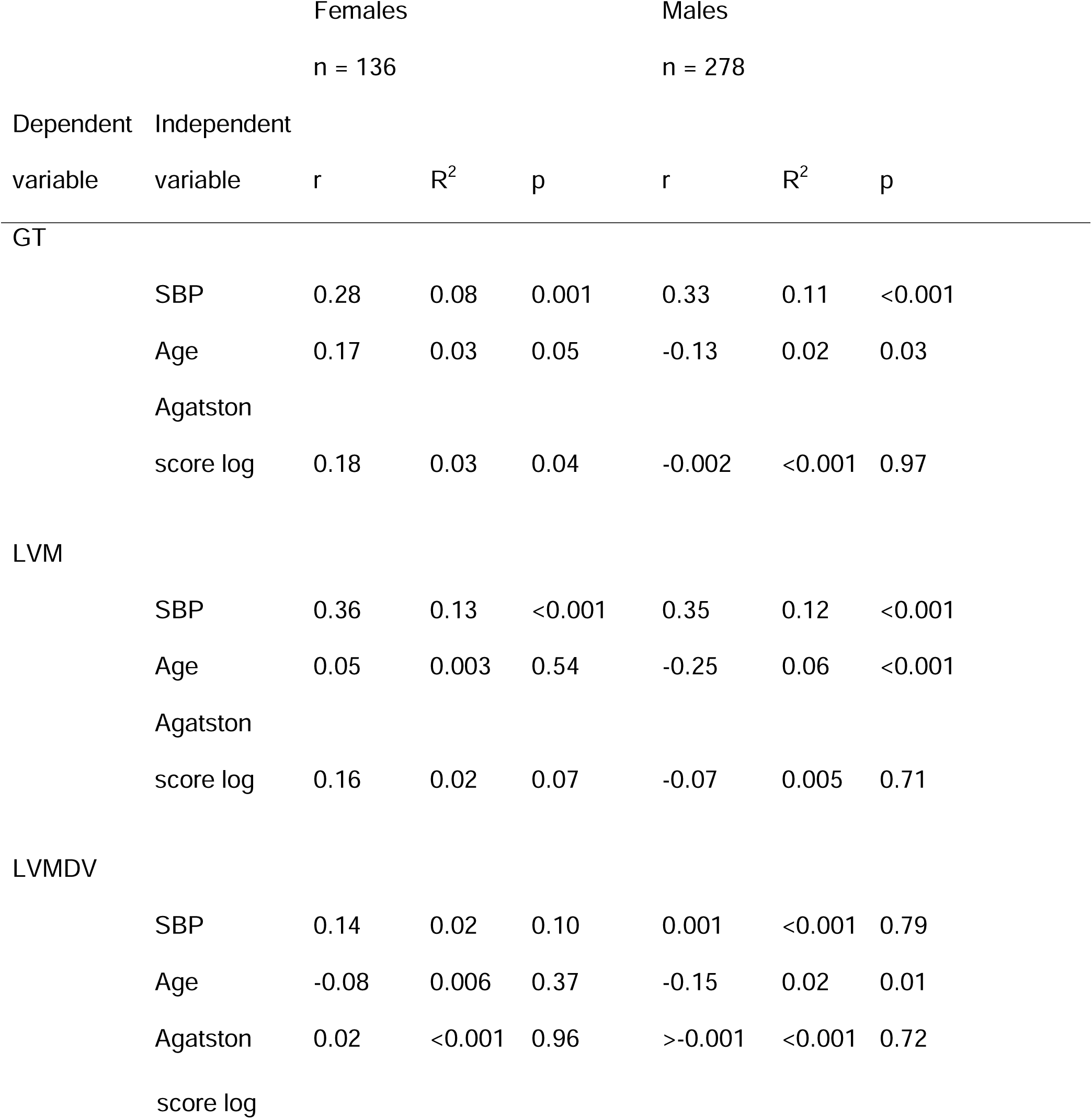
Associations of GT, LV mass and LVMDV by univariable linear regression. Independent values were SBP, age and Agatston score of females (left, n=136) and males (right, n=278). Abbreviations: GT=Global wall thickness, LVM=left ventricle mass, LVMDV=left ventricle mid-diastolic volume, SBP=systolic blood pressure, Agatston score log= Agatston score by logarithmic value.

For males, every mmHg increment in SBP corresponded to a 0.03 mm increase in GT (R^2^=0.11, p<0.001). Neither age nor Agatston score were correlated with GT (p=0.13 and p=0.43, respectively). For females, every mmHg increment in SBP corresponded to a 0.02 mm increase in GT (R^2^=0.08, p=0.001). Age and Agatston score were weakly correlated with GT (R^2^=0.03, p=0.02, and R^2^=0.03, p=0.009, respectively).

For both sexes, LVM increased with increased blood pressure, 0.6 g per mmHg SBP for men, and 0.4 g per mmHg SBP for women. Neither LVM nor LVMDV correlated with Agatston score for both sexes. For men, LVMDV and LVM decreased by 1.17 ml and 1.5 g for every increase in year of age.

## Discussion

The main finding of this study is that GT can be calculated with high accuracy using coronary CTA.

GT is the first method to assess normality of LVM in relation to a given LVEDV, expressed as global wall thickness in mm. Therefore, GT provides an intuitive measure of the balance between left ventricular volume and mass, where the calculated GT has excellent agreement with measured GT of the left ventricle.

Previously, GT has been found to be a strong independent predictor of death and hospitalization for heart failure in patients with normal findings on CMR, including LVEDVi, LVMi and EF within the normal reference ranges. GT appears to be a sensitive measure of early LVH.

The method for estimating GT using only LVM and LVMDV simplifies the method to attain GT, without the need for a software plugin or the need for any particular commercial software for calculations. Sex-specific normal GT reference values were determined to be 5.2-8.8 mm for females and 6.2-11.5 mm for males in this age group of 50-64 years. Calculated GT with coronary CTA formula was similar to GT calculated with the CMR formula and therefore either formula can be used for GT calculation independent of whether coronary CTA or CMR images are available. Normal values for GT at mid-diastole (by coronary CTA) would be expected to be larger than those at end-diastole (by CMR). Indeed, normal values for females were numerically larger (coronary CTA 6.2-11.5 mm vs CMR 4.8-7.1 mm), whereas they were similar for males (coronary CTA 5.2-8.1 mm vs CMR 5.8-8.5 mm). The reason for this inconsistency among males is unclear, but is likely related to the different populations, where the coronary CTA healthy population was on average 20-30 years older, and 4 kg/m^2^ more obese than the CMR healthy population ^13^.

Previous studies have measured LV mean wall thickness as the average of measurements from 17 segments of the LV^13,15,16^, according to American Heart Associatiońs model. Considering that GT by CMR shows improved diagnostic and prognostic performance^17^, it is likely that this is also the case for GT by CT. However, future studies are justified to confirm this assumed prognostic performance. Calculation of GT by CMR is performed using LV measurements in end-diastole. By comparison, CT measurements of the LV are performed in mid-diastole. Therefore, it is anticipated that the GT and blood volume can vary slightly between mid-diastolic measurement on CT compared to end-diastolic measurements by CMR. The method for delineation used in the current study resulted in values that are in accordance with those previously reported for LV volume and mass indexed for body surface area for men and women, obtained from CT examinations.^17^

It is known that higher blood pressure increases LV wall thickness in early stages of disease, but, after some time, causes LV dilation and thinning of the wall.^16^ Wall thinning primarily occurs in the setting of advanced disease, and the diagnostic and prognostic performance of GT by CMR is most prominent in early and less-prominent disease. Other studies have found that older hearts start to show thinner LV wall thickness and a reduction in LVM.^16^ The age group studied in the current study was between 50-64 years of age. Notably, the results show that GT increases modestly with blood pressure, and to small, and likely negligible, extent with age.

The present study evaluated the association between GT and Agatston score, and found either no association (males), or a negligible association (females) between left ventricle GT and different degrees of coronary artery calcification. This suggests that coronary calcification is unrelated to LV wall thickness, and that these two measures, which are both known to be associated with poor prognosis, likely confer independent risk.

## Limitations

Papillary muscles and trabeculations were excluded from the LV mass and included in the blood pool in the current study. Thus, the GT normal values will only be applicable to measurements of LV mass and volume using the same methodology. Furthermore, papillary muscles and trabeculations have been found to confer important complementary diagnostic information in some conditions such as Anderson-Fabry disease^16^, and these changes will not be detected by GT using the proposed methodology.

## Clinical applications

Early detection of cardiovascular disease is important when attempting to fulfill the World Health Organization’s goal for reduced mortality and morbidity. Several types of cardiovascular disease such as heart failure, myocardial ischemia, hypertension, and myocardial interstitial diseases may lead to LVH. This study focuses on a specific way to detect LVH, by measuring the global wall thickness that can be used to categorize LVH. GT adds value and discrimination in LVH amongst those with normal mass (early disease). LV mass is known to be highly prognostic when the mass is elevated (advanced disease). The combination of these two measures can be used to characterize hypertrophy. GT has a prognostic value for hospitalization and death in heart failure, both in an early onset and in advanced disease.^13^ Normal range values of GT using coronary CTA have not previously been determined, and this study has established normal reference ranges of these values in coronary CTA for this age group. These metrics may help identify patients at increased risk for adverse events, supporting the usability for clinical reporting of this variable.

The wide range of values for LV mass and volume in the diseased and healthy groups included in the derivation and validation studies imply that the derived equation should be valid for a wide range of LV sizes, masses and wall thicknesses. Thus, the presented equation can be used for any coronary CTA results where only the values for LV mass and LVMDV are available. If these values are available, no specific software or plugin is needed if these values are available to calculate GT in clinical routine. Regarding sex differences, this study has shown differences in several parameters. However, the same GT formula can be used for both sexes, since the geometric relationship between mass, volume and thickness is not expected to be influenced by sex.

## Conclusion

The current study derived and validated a simple equation for GT based on LV volume and LV mass, and presents sex-specific normal healthy reference ranges for these values using prospectively ECG-triggered coronary CTA.

## Data Availability

All data produced in the present study are available upon reasonable request to the authors.

## Abbreviations

LVH: left ventricular hypertrophy
GT: global wall thickness
LVM: left ventricular mass
CTA: computed tomography angiography
SCAPIS: Swedish CArdioPulmonary bioImage Study
LVMDV: left ventricular mid-diastolic volume
CMR: cardiovascular magnetic resonance
SBP: systolic blood pressure
LV: left ventricle

---

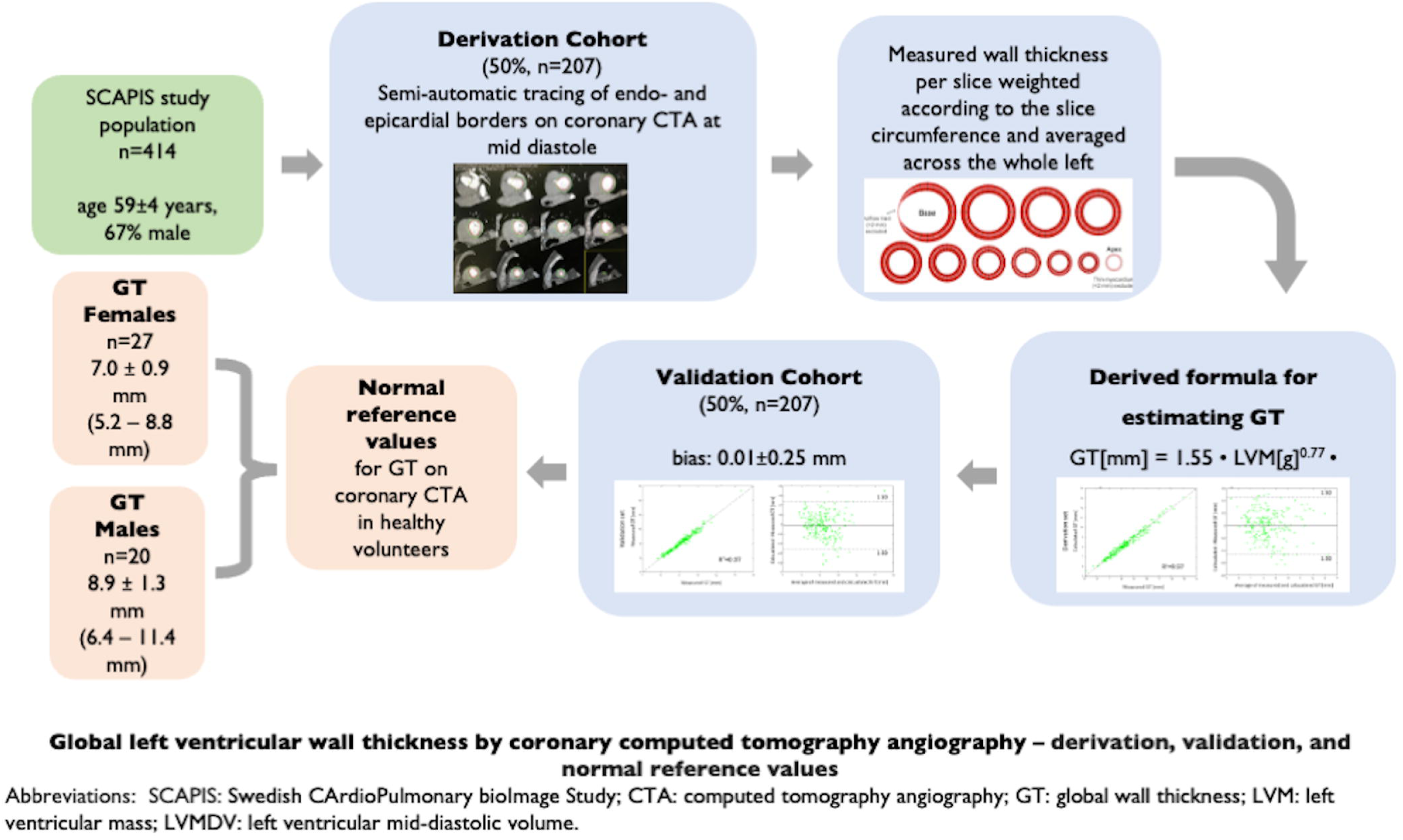

